# Ultra-Processed Food Consumption and Physical, Cognitive and Academic Outcomes in Australian Early Adolescents Aged 10–14 Years: Protocol for a Cross-Sectional Study

**DOI:** 10.64898/2026.11.21.26368181

**Authors:** R. Aysath Shifana, Habisathul Rilha

## Abstract

**Introduction:** Ultra-processed foods (UPFs, NOVA Group 4) provide about half of the daily energy intake of Australian adolescents. Their links with adiposity are well studied. Far less is known about how UPF intake relates to cognition and school achievement in early adolescence, and no Australian study has examined physical, cognitive and academic outcomes together using the NOVA classification.

**Methods and analysis:** This cross-sectional study will recruit approximately 650 students aged 10–14 years from schools sampled across Australian states and territories, stratified by jurisdiction, remoteness and socioeconomic status. Dietary intake will be assessed with two non-consecutive 24-hour recalls (ASA24-Australia), and UPF intake will be expressed as a percentage of total energy. Outcomes are measured BMI z-score, the CogState Brief Battery (Detection, Identification, One Card Learning, One Back) and NAPLAN Reading and Mathematics scores. Multilevel linear regression will account for clustering by school and adjust for socioeconomic position, maternal education, sleep, screen time, physical activity, age and sex. Mediation analysis will explore whether BMI z-score mediates UPF–cognition associations and whether cognition mediates UPF–NAPLAN associations.

**Ethics and dissemination:** Approval will be sought from a university Human Research Ethics Committee and from state, territory, Catholic and independent school authorities before recruitment. Findings will be reported in peer-reviewed journals and at conferences, and summarised for schools, families and policymakers.

**Strengths and limitations of this study:**

- Uses NOVA classification and repeated 24-hour recalls rather than dietary patterns from food frequency questionnaires.
- Combines measured BMI, objective cognitive testing and standardised national achievement data in one sample.
- Samples schools across states and territories, stratified by remoteness and socioeconomic status.
- The cross-sectional design cannot establish temporal order or causation, including for mediation.
- Self-reported dietary recalls are subject to misreporting, and residual confounding is likely.
- NAPLAN results may be one to two years older than the dietary data for some students.

## 1. Introduction

### 1.1 UPF intake in Australian children and adolescents

Ultra-processed foods are industrial formulations of refined ingredients and additives, such as flavourings, colourings and emulsifiers, with little intact whole food. They form Group 4 of the NOVA classification (Monteiro et al., 2019). Typical examples are packaged snacks, sweetened drinks, breakfast cereals, processed meats, instant noodles and ready meals (Levine & Ubbink, 2023).

In the 2011–12 National Nutrition and Physical Activity Survey, UPFs supplied 54.3% of adolescents’ total energy intake (Machado et al., 2020). A more recent study of Victorian schoolchildren aged 8–12 years reported 47.2% (Clark et al., 2025). Intake is shaped by convenience, cost, taste, packaging and marketing (Imtiyaz et al., 2023), and by school food environments and family purchasing. Early adolescents may be especially exposed because they gain freedom over food choices while still depending on home and school.

### 1.2 UPF and physical health

A recent meta-analysis found that higher UPF intake is associated with greater odds of overweight and obesity in adolescents (Aweke et al., 2026). An umbrella review of meta-analyses linked UPF exposure with adverse cardiometabolic outcomes (Lane et al., 2024). In Australian schoolchildren, UPF intake has been related to higher BMI z-scores and central adiposity, though findings vary by age (Clark et al., 2025). Plausible mechanisms include high energy density, palatability, faster eating rates and low fibre and protein content (Temple, 2024). Most of this evidence is observational, and UPF intake is entangled with inactivity, short sleep, screen time and socioeconomic disadvantage.

### 1.3 UPF and cognitive function

Sustained attention continues to develop through adolescence and predicts distractibility in class (Hobbiss & Lavie, 2024). Memory, processing speed and executive function also change markedly in this period. Diets high in UPFs may be energy-rich but low in nutrients important for brain development, such as iron, zinc, B vitamins and essential fatty acids. Proposed pathways include glycaemic fluctuation, systemic inflammation, impaired insulin signalling and oxidative stress (Jacka et al., 2015), and altered gut–brain signalling from low fibre intake. Evidence on food additives and children’s cognition remains limited.

Confounding is a central challenge (Andrade, 2024). Adolescents who eat more UPFs may also sleep less, spend more time on screens, be less active or live with socioeconomic disadvantage, and each of these can independently affect cognitive test results. Maternal education may influence both food availability and support for learning.

### 1.4 UPF and academic performance

In Spanish adolescents, higher UPF consumption was associated with poorer academic performance (López-Gil et al., 2025). Australian research has mostly examined specific food groups, breakfast or overall diet quality rather than total UPF intake (Nyaradi et al., 2013; Pearce et al., 2018). NAPLAN Reading and Mathematics scores provide nationally standardised indicators of achievement and have been used in Australian diet research (Pearce et al., 2018). Achievement is also shaped by prior attainment, attendance, teaching quality, parental support and socioeconomic status.

### 1.5 Evidence gap

Australian evidence on diet, cognition and achievement comes largely from the Western Australian Pregnancy Cohort (Raine) Study. A Western dietary pattern high in takeaway foods, confectionery and soft drinks was associated with poorer cognitive outcomes and academic performance (Nyaradi et al., 2014, 2015). Among 315 children aged 9–11 years from 26 schools, a non-core dietary pattern was associated with lower NAPLAN scores, a difference of 12.6 points that attenuated to 8.5 points after adjustment (Pearce et al., 2018).

These studies have three limitations. They derived population-specific dietary patterns from food frequency questionnaires rather than classifying foods by degree of processing. Their data are now more than a decade old. They did not quantify UPF exposure with standardised 24-hour recall tools such as ASA24 (Subar et al., 2012). No Australian study has examined NOVA-defined UPF intake alongside measured BMI, objective cognitive testing and NAPLAN results in a contemporary sample of early adolescents.

### 1.6 Aim and research questions

The study aims to examine associations between UPF intake and physical, cognitive and academic outcomes in Australian adolescents aged 10–14 years, after adjustment for relevant confounders.

- **RQ1:** Is UPF intake (% of total energy) associated with BMI z-score?
- **RQ2:** Is UPF intake associated with cognitive performance on the CogState Brief Battery (Detection, Identification, One Card Learning, One Back)?
- **RQ3:** Does BMI z-score mediate the association between UPF intake and cognitive performance?
- **RQ4:** Is UPF intake associated with NAPLAN Reading and Mathematics scores?
- **RQ5:** Does cognitive performance mediate the association between UPF intake and NAPLAN scores?

We hypothesise that higher UPF intake will be associated with higher BMI z-scores, poorer cognitive performance and lower NAPLAN scores.

## Methods and analysis

### 2.1 Study design

This is a quantitative, cross-sectional observational study. Dietary intake, anthropometry, cognitive function, academic performance and confounders will be measured at a single time point. The design allows several associations to be examined together but cannot establish temporal order or causation (Li et al., 2025). The study will be reported following the STROBE guidelines for cross-sectional studies.

### 2.2 Setting, participants and sampling

Schools across Australian states and territories will be stratified by jurisdiction, geographic remoteness and socioeconomic status. Schools, and then students within schools, will be randomly selected to obtain a representative sample. Students are eligible if they are aged 10– 14 years, attend a participating school, give their own assent and have written parental or guardian consent. Students unable to complete the dietary recall or computerised cognitive tasks, even with reasonable adjustments, will be excluded.

### 2.3 Sample size

The target sample is approximately 650 students. Detecting a small effect (f^2^ = 0.02) for a single exposure coefficient in a linear regression model with ten predictors, at a two-sided α of

0.05 and 80% power, requires 395 participants. Allowing for clustering of about 25 students per school with an intraclass correlation of 0.02 (design effect 1.48) raises this to 585. Inflating by 10% for incomplete data gives approximately 650.

**Table 1.** Study variables and measurement.

| **Variable** | **Measurement** |
|---|---|
| UPF intake (exposure) | Two non-consecutive 24-hour dietary recalls using ASA24-Australia (Subar et al., 2012; Clark et al., 2025). Foods classified with NOVA; UPF intake expressed as % of total energy (Monteiro et al., 2019). |
| Physical health | Height and weight measured privately by trained staff; BMI-for-age z-scores calculated with WHO AnthroPlus. |
| Cognitive function | CogState Brief Battery: psychomotor speed (Detection), attention (Identification), visual learning (One Card Learning) and working memory (One Back). |
| Academic performance | NAPLAN Reading and Mathematics scale scores (Year 5, 7 or 9), obtained with parent or guardian consent from schools or the relevant test administration authority. |
| Confounders | Area-level socioeconomic position (SEIFA), maternal education, sleep duration, screen time, physical activity, age and sex. |

### 2.4 Measures

Because NAPLAN is taken in Years 5, 7 and 9, year level and time since testing will be recorded and accounted for in analyses. Foods will be assigned to NOVA groups by two coders working independently, with disagreements resolved by discussion.

### 2.5 Data management

Participants will be identified by study codes rather than names. Electronic files will be encrypted and stored on secure university servers, with access limited to authorised members of the research team.

### 2.6 Statistical analysis

All analyses will be conducted in R. Descriptive statistics will summarise the sample, UPF intake and outcomes. Associations between UPF intake and each outcome (RQ1, RQ2, RQ4) will be estimated with multilevel linear regression, with students nested in schools and, where appropriate, schools in states or territories. Models will adjust for the confounders listed above. Cognitive and academic outcomes will first be analysed separately.

Mediation analysis will then examine indirect pathways via BMI z-score (RQ3) and cognitive performance (RQ5). Because the data are cross-sectional, indirect effects will be interpreted as statistical associations consistent with mediation, not as evidence of causal pathways. Missing data will be described and, where appropriate, handled with multiple imputation. Statistical significance will be set at a two-sided α of 0.05.

**Table 2.** Study timeline.

| **Period** | **Activities** |
|---|---|
| Year 1 | Finalise protocol and literature review; ethics and school authority approvals |
| Year 2 | Recruit schools and participants; collect dietary, anthropometric, cognitive and NAPLAN data |
| Year 3 | Clean and analyse data; write up and disseminate findings |

### 2.7 Timeline

## Data Availability

Not applicable. This is a study protocol; no data have been collected.

https://orcid.org/0009-0007-7856-6222

## 3. Ethics and dissemination

Ethics approval has not yet been obtained. Approval will be sought from a university Human Research Ethics Committee, in line with the National Statement on Ethical Conduct in Human Research (National Health and Medical Research Council et al., 2025). Approval will also be sought from state and territory education departments, Catholic and independent school authorities, and each participating school before recruitment.

Written parental or guardian permission and adolescent assent will be obtained, with age-appropriate information (Brothers et al., 2020). Participation is voluntary, and students may withdraw at any time without any effect on their schooling. Access to NAPLAN results requires explicit parental consent and approval from the relevant test administration authority. Height and weight will be measured privately to minimise embarrassment and weight stigma. Referral support will be offered if participation raises concerns about nutrition or wellbeing.

Findings will be published in peer-reviewed journals and presented at conferences. Plain-language summaries will be prepared for participating schools, families and policymakers, with relevance to school canteen, food labelling and marketing policy.

## Declarations

### Author contributions

A.S.R. conceived the study, designed the protocol and wrote the manuscript. H.R. provided methodological guidance and critically revised the manuscript. Both authors approved the final version.

### Data availability

Not applicable; no data have been collected.

### Use of AI tools

Claude (Anthropic) was used to restructure and edit the manuscript. The authors reviewed all content and take full responsibility for it.

